# Subnational analysis of the COVID-19 epidemic in Brazil

**DOI:** 10.1101/2020.05.09.20096701

**Authors:** Thomas A Mellan, Henrique H Hoeltgebaum, Swapnil Mishra, Charlie Whittaker, Ricardo P Schnekenberg, Axel Gandy, H Juliette T Unwin, Michaela A C Vollmer, Helen Coupland, Iwona Hawryluk, Nuno Rodrigues Faria, Juan Vesga, Harrison Zhu, Michael Hutchinson, Oliver Ratmann, Mélodie Monod, Kylie E C Ainslie, Marc Baguelin, Sangeeta Bhatia, Adhiratha Boonyasiri, Nicholas Brazeau, Giovanni Charles, Zulma Cucunuba, Gina Cuomo-Dannenburg, Amy Dighe, Jeff Eaton, Sabine L van Elsland, Katy A M Gaythorpe, Will Green, Edward Knock, Daniel Laydon, John A Lees, Andria Mousa, Gemma Nedjati-Gilani, Pierre Nouvellet, Kris V Parag, Hayley A Thompson, Robert Verity, Caroline E Walters, Haowei Wang, Yuanrong Wang, Oliver J Watson, Lilith Whittles, Xiaoyue Xi, Ilaria Dorigatti, Patrick Walker, Azra C Ghani, Steven Riley, Neil M Ferguson, Christl A Donnelly, Seth Flaxman, Samir Bhatt

## Abstract

Brazil is currently reporting the second highest number of COVID-19 deaths in the world. Here we characterise the initial dynamics of COVID-19 across the country and assess the impact of non-pharmaceutical interventions (NPIs) that were implemented using a semi-mechanistic Bayesian hierarchical modelling approach. Our results highlight the significant impact these NPIs had across states, reducing an average *R_t_ >* 3 to an average of 1.5 by 9-May-2020, but that these interventions failed to reduce *R_t_* < 1, congruent with the worsening epidemic Brazil has experienced since. We identify extensive heterogeneity in the epidemic trajectory across Brazil, with the estimated number of days to reach 0.1% of the state population infected since the first nationally recorded case ranging from 20 days in São Paulo compared to 60 days in Goiás, underscoring the importance of sub-national analyses in understanding asynchronous state-level epidemics underlying the national spread and burden of COVID-19.

## 2 Introduction

The world faces an unprecedented public health emergency in the COVID-19 pandemic. Since the emergence of the novel coronavirus (SARS-CoV-2) in China in December 2019, global spread has been rapid, with over 10 million cases and over 500 thousand deaths reported globally as of the 30th June [18]. Though transmission of the disease beyond Asia was initially centred around Western Europe and North America, significant spread is now seen in other parts of the world, including many countries across Sub-Saharan Africa [16, 15, 17] and Latin America [14, 2].

One such area is Brazil - since report of its first case on 26th February, its epidemic has grown quickly, with the country now reporting over 1,600,000 cases and over 65,000 deaths [18]. In response to significant spread and community transmission of the virus within the country, Brazilian state and city officials implemented extensive public health measures to reduce the transmission of COVID-19, including declaring a state of emergency, mandating the closure of retail and service businesses, restricting transportation, and closing schools. Specific packages of interventions have been decided at the state level, with substantial variation between states in their comparative timing and the extent to which measures have been adopted [3]. Importantly, the interventions employed remained short of the widespread and mandatory lockdowns implemented across parts of Asia and Europe which have proved to be highly effective at containing spread of the virus [7, 25].

Given current evidence for sustained circulation of the virus in Brazil - reported daily deaths regularly exceed 1000 individuals per day [19] - a better understanding of the epidemiological origin and the impact of those initial interventions deployed is required to guide current policy decisions aimed at preventing worsening of the public health emergency the country faces. Motivated by this, we extend a previously published semi-mechanistic Bayesian hierarchical model of COVID-19 epidemiological dynamics [7, 25] to assess the impact of interventions aimed at curbing transmission of COVID-19 across Brazil. In our framework we estimate the number of deaths, infections and transmission as a function of patterns in human mobility. We utilise this framework to explore the epidemiological situation in detail at the state level, understand the highly heterogeneous spread of the virus across the country and understand the insufficiency of the initial response to the virus across states.

## 3 Results

Across the 18 Brazilian states with more than 50 deaths over the time period up to 9th May, we estimate that implemented NPIs had a substantial impact on transmission and spread of SARS-CoV-2, reducing *R_t_* from greater than three to 1.5 (95% CI 1.3-1.9) on average. Figure 1 shows the model estimated time-varying reproduction number (*R_t_*) for 5 states - São Paulo (SP), Rio de Janeiro (RJ), Amazonas (AM), Maranhão (MA) and Bahia (BA) - chosen to reflect the extensive variation in the trajectory and evolution of the epidemic observed between states (although see Supplementary Figure 1 for analyses of all 18 states that at the date considered here had more than 50 deaths total). São Paulo and Rio de Janeiro have the highest numbers of deaths, Amazonas has the highest predicted attack rates, while the epidemics in Maranhão and Bahia are comparatively nascent in progression. In each of the five states estimates of the initial reproduction number (*R*_0_) prior to intervention are consistently in the range of 3 - 4, in line with estimated values of transmissibility derived from European data [7] - this was observed across all states analysed here except for Amapa (AP) and Alagoas (AL) where interventions were implemented early relative to the first recorded COVID-19 deaths in these states, resulting in estimated *R*_0_ values near 2. Across all states, our results highlight that *R_t_* has dropped consistently following the implementation of public health interventions. For the 5 states considered in Figure 1, mobility indicators dropped by 33% on average by May 9th across Brazil compared to a baseline derived from data on the same date in the preceding year, and *R_t_* declined by greater than 50% on average, for example to values of 0.8 (95% CI: 0.6%-1.0%) in Maranhão and 1.6 (95% CI: 1.3%-1.9%) in Bahia. Despite these substantial reductions in *R_t_* however, our results also indicate it is unlikely that these measures have brought *R_t_* consistently below 1. This was observed across all states analysed and highlights the insufficiency of the initial NPIs implemented at a state level in controlling and preventing further growth of the COVID-19 epidemic.

**Figure 1:**
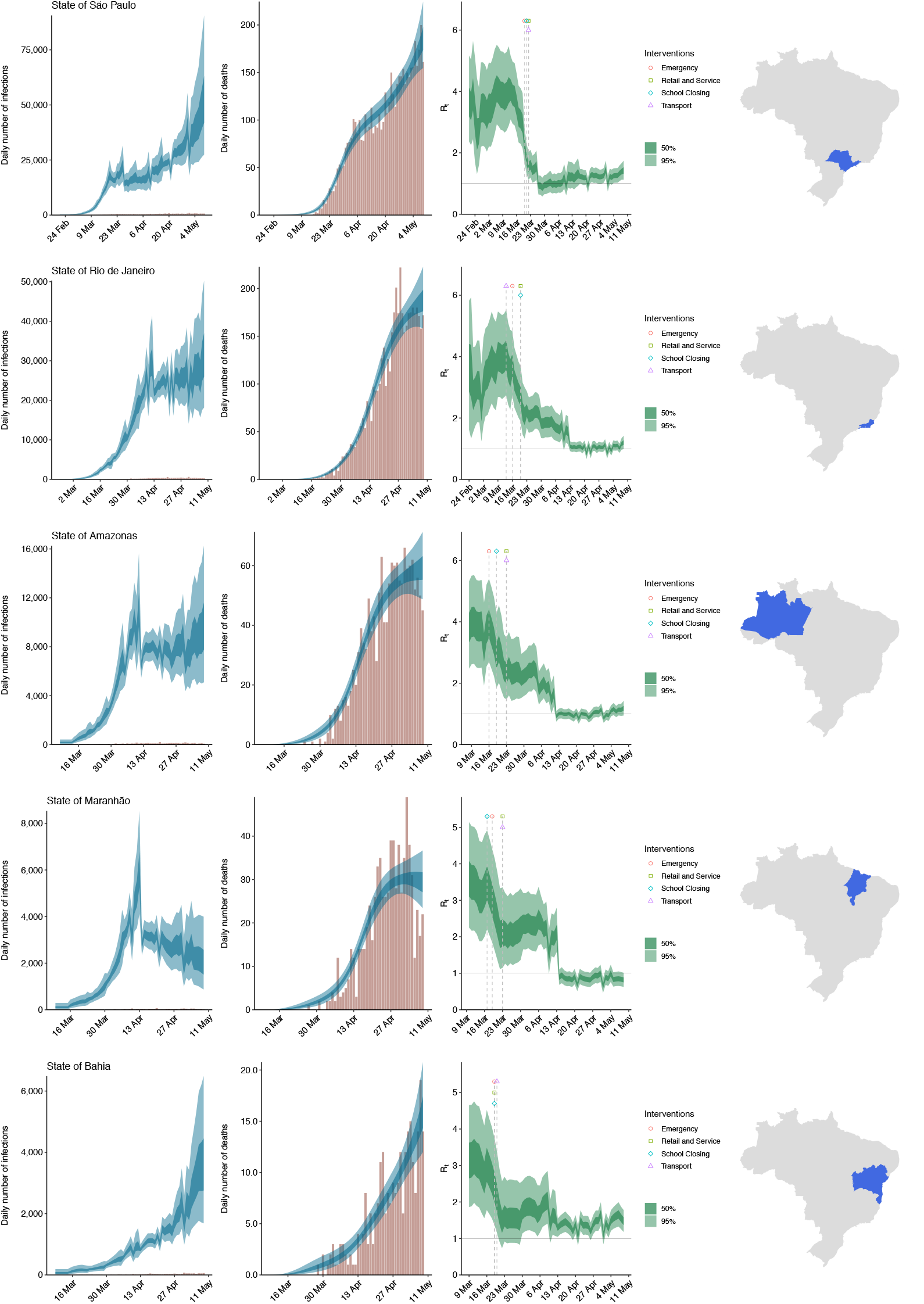
Estimates of infections, deaths and *R_t_* for selected states. Left: daily number of infections, brown bars are reported cases, blue bands are predicted infections, dark blue 50% credible interval (CI), light blue 95% CI. Middle: daily number of deaths, brown bars are reported deaths, blue bands are predicted deaths, CI as in left plot. Right: time-varying reproduction number *R_t_*, dark green 50% CI, light green 95% CI. If the *R_t_* is above 1, the number of infections continues to grow. Icons are interventions shown at the time they occurred.

Despite predicted similarities between states in estimates of *R*_0_ and the impact of implemented NPIs, there have been striking asymmetries in the burden of COVID-19 experienced across the country over the time period. For example the two states of São Paulo and Rio de Janeiro alone account for greater than 50% of recorded deaths out of the 18 states considered in our model. Our analyses similarly support profound geographical variation in the dynamics and burden of COVID-19 across Brazil. In terms of epidemic timing relative to reports of the first confirmed COVID-19 case in Brazil (recorded in São Paulo on the 26th-Feb), our results suggest substantial variation between states in the timing of virus establishment and subsequent community transmission (assessed here as the time taken for each state to reach a predicted attack rate of 0.1% of the population, Figure 2). This ranged from only 20 days (95% CI: 18-21) in São Paulo through to greater than 40 days with 95% credibility in Bahia, Rio Grande do Sul, Goiás and Minas Gerais. These results highlight extensive geographical variation in the evolution of epidemic trajectories between states to date, a feature likely attributable to a combination of differences in the timing of state-specific seeding in conjunction with variation in the state-specific transmissibility of the virus (as evidenced by the variation in the baseline *R*_0_ and subsequent reduction in *R_t_* observed between states). Intriguingly, the comparative timing of virus establishment and takeoff in these states lacked a clear geographical pattern, with both states in the South and North of the country showing evidence of early and rapid epidemic takeoff, in contrast to states in the Central-West region of the country where initial seeding appears to have occurred later.

**Figure 2:**
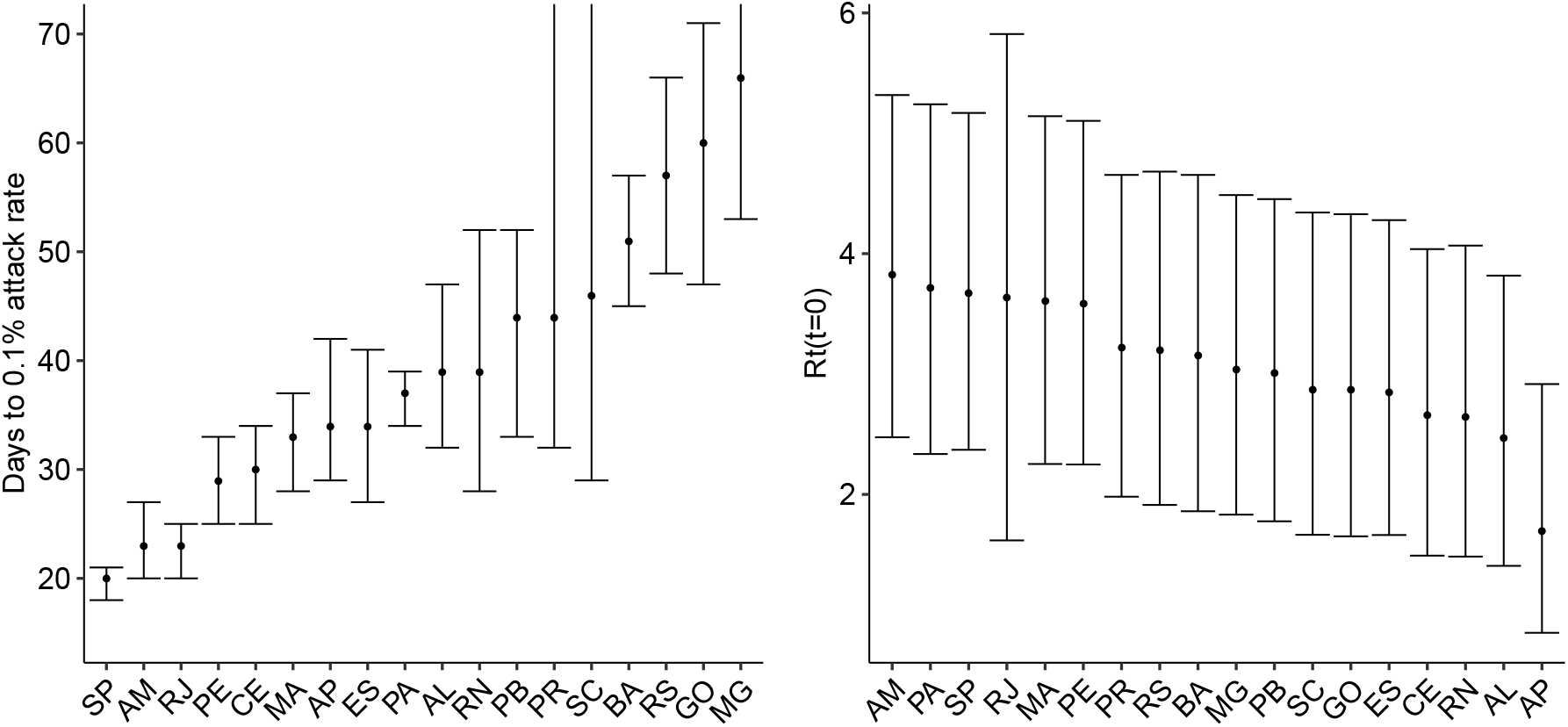
Time in days to 0.1% attack rate (measured from date of first case in Brazil), and initial reproduction number *R_t_*(*t* = 0) across different states: São Paulo (SP), Rio de Janeiro (RJ), Pará (PA), Ceará (CE), Amazonas (AM), Pernambuco (PE), Maranhão (MA), Bahia (BA), Espírito Santo (ES), Alagoas (AL), Paráíba (PB), Minas Gerais (MG), Paráná (PR), Rio Grande do Sul (RS), Rio Grande do Norte (RN), Amapa (AP), Santa Catarina (SC), Goias (GO). Note, RS, GO and PR states had not reached with 0.1% attack rate with 95% CI within the epidemic time analysed.

Similar variation was observed when the predicted attack rates (the total proportion of the population infected over the time period considered) were assessed, with our analyses suggesting attack rates as high as 8% in Amazonas to as low as 0.1% in Minas Gerais (Table 1). Despite this heterogeneity however, estimates across all 18 states suggest that the proportion of the population in each state infected up to the 9th May 2020 was low and short of the herd immunity threshold required to prevent rapid resurgence of the virus if control measures are fully relaxed. These results are driven in part by modelling assumptions regarding the extent of death underreporting and the assumed state-specific IFR and we therefore undertook a series of sensitivity analyses (see Supplementary information 4) exploring different assumptions surrounding state-level IFR (relating to assumptions about how healthcare quality varies with state income) and the extent of death underreporting. The results of these sensitivity analysis yield quantitative differences in the predicted attack rates - for example, assuming a 50% level of death underreporting changes our predicted attack rates from 3.0% (95% CI: 2.1%-4.1%) and 8.0% (95% CI: 5.7%-10.8%) to 6.1% (CI95: 3.9%-9.4%) and 16.5% (CI95:3.8%-25.4%) for São Paulo and Amazonas respectively. Varying assumptions surrounding the extent and variation of healthcare quality across states similarly altered the estimates of predicted attack rates, but did not qualitatively alter any of the conclusions reached, namely that the observed levels of infection in the population are significantly lower than that required for herd immunity.

**Table 1:** Estimated infection fatality ratio (IFR), state population, reported deaths and deaths per million population, estimated number of infections in thousands, attack rate (AR), and time-varying reproduction number on 9th-May-2020 with 95% credible intervals, for São Paulo (SP), Rio de Janeiro (RJ), Pará (PA), Ceará (CE), Amazonas (AM), Pernambuco (PE), Maranhão (MA), Bahia (BA), Espírito Santo (ES), Alagoas (AL), Paráíba (PB), Minas Gerais (MG), Paráná (PR), Rio Grande do Sul (RS), Rio Grande do Norte (RN), Amapa (AP), Santa Catarina (SC), Goias (GO).

| State | IFR % | Population | Deaths | Deaths per million | Infections (thousands) | Attack rate % | $R_t$ |
| --- | --- | --- | --- | --- | --- | --- | --- |
| SP | 0.7 | 46,289,333 | 5,142 | 111 | 1,370,000 [950,000, 1,880,000] | 3.0 [2.1, 4.1] | 1.4 [1.1, 1.7] |
| RJ | 0.8 | 17,366,189 | 4,564 | 263 | 1,050,000 [777,000, 1,380,000] | 6.1 [4.5, 8.0] | 1.1 [0.9, 1.3] |
| PA | 0.9 | 8,690,745 | 1,752 | 202 | 529,000 [376,000, 724,000] | 6.1 [4.3, 8.3] | 1.3 [1.1, 1.5] |
| CE | 1.1 | 9,187,886 | 1,573 | 171 | 508,000 [381,000, 661,000] | 5.5 [4.1, 7.2] | 1.7 [1.5, 1.8] |
| AM | 0.8 | 4,207,714 | 1,446 | 344 | 337,000 [241,000, 454,000] | 8.0 [5.7, 10.8] | 1.2 [0.9, 1.4] |
| PE | 1.1 | 9,617,072 | 1,422 | 148 | 290,000 [203,000, 400,000] | 3.0 [2.1, 4.2] | 1.4 [1.1, 1.6] |
| MA | 1.0 | 7,114,598 | 745 | 105 | 121,000 [81,800, 172,000] | 1.7 [1.2, 2.4] | 0.8 [0.6, 1.0] |
| BA | 1.1 | 14,930,424 | 263 | 17 | 62,800 [39,100, 93,800] | 0.4 [0.3, 0.6] | 1.6 [1.3, 1.9] |
| ES | 0.9 | 4,064,052 | 202 | 50 | 57,100 [33,700, 88,500] | 1.4 [0.8, 2.2] | 1.5 [1.1, 1.8] |
| AL | 1.1 | 3,351,092 | 182 | 54 | 65,700 [37,700, 103,000] | 2.0 [1.1, 3.1] | 1.8 [1.4, 2.3] |
| PB | 1.2 | 4,039,277 | 161 | 40 | 58,900 [35,000, 92,300] | 1.5 [0.9, 2.3] | 2.1 [1.7, 2.5] |
| MG | 1.0 | 21,292,666 | 153 | 7 | 30,700 [15,200, 53,700] | 0.1 [0.1, 0.3] | 1.5 [1.0, 2.0] |
| PR | 0.9 | 11,516,840 | 113 | 10 | 20,800 [9,980, 36,800] | 0.2 [0.1, 0.3] | 1.2 [0.7, 1.7] |
| RS | 0.9 | 11,422,973 | 106 | 9 | 35,200 [17,100, 62,700] | 0.3 [0.2, 0.6] | 1.8 [1.3, 2.4] |
| RN | 1.1 | 3,534,165 | 90 | 26 | 21,400 [10,200, 38,400] | 0.6 [0.3, 1.1] | 1.6 [1.1, 2.1] |
| AP | 0.7 | 861,773 | 69 | 80 | 31,000 [15,000, 54,600] | 3.6 [1.7, 6.3] | 1.6 [1.2, 2.0] |
| SC | 0.8 | 7,252,502 | 68 | 9 | 16,800 [6,850, 32,500] | 0.2 [0.1, 0.5] | 1.4 [0.9, 2.0] |
| GO | 0.9 | 7,116,143 | 60 | 8 | 21,600 [8,870, 42,300] | 0.3 [0.1, 0.6] | 2.0 [1.4, 2.6] |

## 4 Discussion

Attempts to contain the spread of SARS-CoV-2 in the community have centred around the deployment of various non-pharmaceutical interventions (NPIs) that involve reducing the number of contacts made between individuals [6], in doing so attempting to disrupt chains of transmission, bring the reproduction number (*R_t_*) below 1 and curb exponential growth of the epidemic. Common examples of these NPIs include school closures, social distancing rules, banning of public gatherings and complete lockdown, which together have been shown to be effective in reducing *R_t_* across a number of different settings [7]. Using our framework, with *R_t_* parameterised as a function of Google mobility data [1], we highlight the marked effect of NPIs on transmission in Brazil over the period considered here, which substantially slowed spread of the virus. Despite these reductions however, our results also highlight that reductions in mobility (which saw a 33% reduction relative to baseline) were not stringent enough to reduce *R_t_* below 1 in many states. This is consistent with the interventions implemented in Brazil over the period - whilst there was substantial variation between states in the exact measures adopted, all interventions remained short of the wide-reaching lockdowns implemented across, for example, parts of Europe and which have effectively controlled transmission of the virus. In previously published work examining Italy [25] where stringent measures including societal lockdowns have been implemented, mobility reduced by 53% compared to baseline [1], a substantially larger decrease than that observed across Brazilian states. These reductions in movement across Italy were predicted to have reduced *R_t_* by 85% compared to *R*_0_, bringing it significantly below 1. This is in contrast to Brazil where we estimate reductions in mobility have only reduced *R_t_* by 55% on average, making it unlikely that *R_t_* has decreased below 1, a result supported by the worsening of the epidemic in Brazil in the weeks following our cutoff date.

Brazil has already reported more than 1,600,000 cases and 65,000 deaths,[18] more than ten times as many COVID-19 deaths and cases as China. Despite these high numbers for the country as a whole, there have been noticeable differences in the burden of COVID-19 experienced between states to date, with the distribution of deaths among states highly heterogeneous. Our results support this geographical variation, highlighting specifically the variation in the likely timing of epidemic takeoff that has led to the asynchronous epidemics that have emerged across different states. Both this and the observed lack of geographical contiguity in the timing of takeoff between states is consistent with recent work emphasising the initial local spread of the virus within states where seeding from international sources had occurred (e.g. the major urban hubs of São Paulo and Rio de Janeiro) before national air travel ferried the virus and established transmission in the rest of the country [5]. This variation highlights that as of the 9th May, Brazilian states were at very different points in their respective epidemics - whereas some had fully established epidemics such as the Amazonas region, others, such as Bahia and Rio Grande do Sul, had only nascent emerging epidemics. Such differences have material consequences for the likely evolution of the epidemic trajectories by state in the coming months. Moreover, they underscore the importance of granular, sub-national analyses in understanding the spread of SARS-CoV-2, revealing a level of heterogeneity that would be obscured by analyses conducted at the national level.

Despite estimates of *R*_0_ broadly in line with that observed for European settings (i.e. in the range 2.5-4), we also observed heterogeneity in the transmissibility of the virus for each state, with *R*_0_ varying from as high as 3.8 (95% CI: 2.5-5.3) in the Amazonas region to 2.5 (95% CI: 1.4-3.8) in Alagoas (excluding the state of Amapa where the estimated *R*_0_ is lower due to early intervention relative to the first reported cases). Such variation is consistent with patterns of growth observed globally,[8] a phenomenon likely attributable to a variety of factors including the demography of the population (and associated comparative susceptibilities of different age-groups[4]), the spatial structure of urban centres[20] and the baseline level of various individual behaviours (such as mask wearing) associated with reducing transmission of the virus[27]. Despite this variation, our results underscore the comparatively small proportion of the population infected to date, even in the worst affected states, a result congruent with recent serological evidence from a nationwide survey of Brazilian cities [10]. Considering an *R*_0_ of 3, the estimated share of the population infected remains far short of the approximate 70% herd immunity threshold required to prevent resurgence of the virus as control measures are fully relaxed across many states.

Substantial uncertainty remains both in our understanding of the fundamental epidemiology of COVID-19 and the quality of surveillance systems across different settings. Consequently, estimates of the expected number of deaths and infections are sensitive to assumptions made within our modelling framework. In particular there is uncertainty surrounding the infection fatality ratio (IFR), which is the probability of an individual dying if infected with SARS-CoV-2 and which is driven by a complex interplay of factors including population demography and healthcare capacity/quality [26] amongst others. There is also considerable uncertainty in the observed death data, and how patterns of death underreporting vary across both time and space. Despite these uncertainties however, the qualitative conclusions reached here about infection levels being short of the herd immunity threshold were robust to our assumptions about both the state-specific IFR and the degree of death underreporting assumed. Although differences in these assumptions alter our quantitative estimates of attack rate, they do not alter our conclusions surrounding the impact of control interventions and the proportion of the population infected up to the period ending 9th May - namely that it falls far short of the threshold required for herd immunity.

Overall, our results reveal that despite extensive spread of SARS-CoV-2 across Brazil, and despite marked variation between states in the extent of the initial epidemic experienced over the period up to 9th May, the extent of infection in the general population was low and far short of the level required for herd immunity. This result is robust to assumptions surrounding the IFR associated with each state, and the extent of underreporting we assume in the available deaths data. Moreover, whilst our results suggest substantial reductions in the estimated value of *R_t_* across all states following introduction of NPIs, they also underscore the insufficiency of these interventions in bringing *R_t_* below 1 and that without introduction of further control measures, Brazil faces the prospect of an epidemic that will continue to grow.

## 5 Methods

Data: Our model utilises daily consolidated deaths data at the state level to infer epidemiological characteristics of viral spread and transmission to date. The analyses presented here are based on daily death figures, disaggregated by state, from two sources that are published by the Brazilian Ministry of Health. These are *Painel Coronavírus* [19], which is the source for official daily updates on deaths by date of notification to the government. The other is *SIVEP-Gripe* [22], which is updated weekly and provides death counts by the actual date of death. There are limitations associated with both of these datasets highlighted in Supplementary Figure 8 - namely right-hand censoring of recent deaths (e.g. due to delays in testing results such that a COVID-19 diagnosis is only firmly established post-mortem, leading to a delay in inclusion of that death in the database) and/or underreporting of deaths (resulting in the absence of a death from ever featuring in either of the databases).

We employ a variety of strategies to mitigate these issues. To account for delays in death inclusion, we utilise datasets from the 16th June 2020, but restrict our analyses to the period ending 9th May 2020. This is sufficiently long after NPIs were implemented across Brazil for us to draw conclusions about their effectiveness, but sufficiently far back in time to minimise the potential impact of delays in test confirmation and reporting of deaths that results in right-censoring of the data. To account for under-reporting of deaths (i.e. deaths that either occur in the community or that occur in a hospital setting but fail to be recorded), we employ two strategies. Firstly, within our modelling framework, we use the dataset that for a given state maximises the number of cumulative deaths recorded to date, as a strategy to choose the dataset with fewest number of cumulative deaths missing. This accounts for potential between-dataset differences in the time-varying quality of death reporting. In order to account for deaths that are absent from both datasets, we also undertake a series of sensitivity analysis assuming different levels of death underreporting across states (see Supplementary Information for further details).

### Model

In this modelling study, we adapt and extend a previously published Bayesian semi-mechanistic model of COVID-19 transmission and mortality [7], updating the model to explicitly incorporate population level metrics of mobility (specifically, Google Mobility data) that have previously been shown to reflect patterns of transmission across a variety of settings [23] and a weekly autoregressive process intended to capture variation in patterns of transmission between states above and beyond that reflected in their comparative patterns of mobility (e.g. that could reflect variation in adoption of individual-level behaviours that would modify the impact of mobility reductions on transmission). Parameters governing the model are jointly estimated for 18 Brazilian states to evaluate the impact of control interventions on SARS-CoV-2 transmission and to explore variation in epidemic trajectories between states. Detailed descriptions of the model are available elsewhere and code for this model are available at https://github.com/ImperialCollegeLondon/covid19model, but key elaborations and extensions included here are described below.

The time-varying reproduction number *R_t,m_* is estimated at the state level for Brazil, using a similar modelling approach to that utilised in [23]. Specifically the functional form for *R_t_,_m_* is specified in the following way:

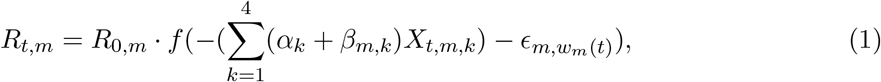

with *f*(*x*) = 2 exp(*x*)/(1 + exp(*x*)) denoting the logistic function multiplied by two, where m indexes each states and where *k* indexes the four different Google Mobility covariates used here, which are Retail & Recreation, Grocery & Pharmacy, Workplaces and Residential and where *α_k_* is a covariate linking each of these mobility metrics to transmission that is shared across states, and *β_m_,_k_* is an additional covariate allowing a state-specific relationship between a particular mobility covariate and transmission. Prior distributions on the covariates were specified as follows:

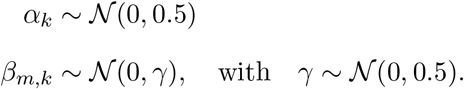

where the variable *∊_m_,_Wm_*(*_t_*) is a weekly auto regressive process with two lags (AR(2)), centred around 0, which intend to capture extra variation between states that is not fully explained by mobility alone. For a full detailed mathematical description of the AR(2) process, see Section 5.1 from [23].

In addition to this elaboration to the formulation of *R_t_*, we utilised Brazil-specific estimates of the key parameters governing the model. The distribution of times from infection to death (infection-to-death) was estimated using patient level data from the 16th June 2020 SIVEP-Gripe dataset[22], specifically through fitting a gamma distribution to the data using a Bayesian MCMC-based framework implemented in the probabilistic programming language STAN. Based on these results, we model the infection-to-death distribution n as a sum of the infection-to-symptom and symptom-to-death distributions,

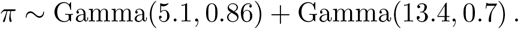

### IFR Calculation

In order to derive an expected IFR across different states in Brazil, mixing patterns from Latin America [9] were integrated with recent reports of disease severity original derived from the Chinese epidemic [21] and subsequently modified to match data from the outbreak in UK [7]. In order to further refine these estimates for a Brazilian setting, and in light of the substantial evidence pointing to substantial increasing risk of COVID-19 mortality with age, we first adjusted estimates of the IFR to the demographic composition of the Brazilian population. These adjusted estimates better reflect the likely severity of COVID-19 in Brazil, but are still predicated on assumptions on comparable quality of healthcare between Brazilian states and the UK. Recent work has highlighted the extent to which 1) variation in healthcare quality can impact the likely IFR in a given setting and 2) how this variation in healthcare quality likely varies with the comparative affluence of a region [26] - we therefore adapted our IFR estimates for each state in an income-dependent manner.

Across the states considered in this analysis, average income (in US dollars) varies from as high as ~ $300 in São Paulo to as low as ~ $100 in Maranhão.[12] Such disparities in income are likely to result in significant differences in the quality and extent of available healthcare. We modified the state-specific IFRs in an income-dependent manner - specifically, we assumed that the state with the highest income (São Paulo) has a quality of care identical to that observed in the UK (and thus motivated using the estimates presented in Verity et al.[21]), and that the state with the lowest income (Maranhão) had significantly worse healthcare outcomes - more similar to those that would be expected in a Lower Middle Income Country (see [26] for further details on how differences in health quality across settings are likely to impact outcomes). For the other states where income lies somewhere between that of Maranhão and São Paulo, we linearly interpolate the age-specific infection fatality probabilities based on state-level average income.[12] These age-specific infection fatality probabilities are then combined with predictions of the age-distribution of infections to produce an overall, state-specific IFR. Because of the inherent uncertainty associated with these modifications, we undertake a number of sensitivity analyses examining how IFR-related assumptions qualitatively impact our results.

## Data Availability

All data and code used is publicly available.

https://covid.saude.gov.br/

https://www.ibge.gov.br/

https://github.com/ImperialCollegeLondon/covid19model

## Acknowledgements

This work was supported by Centre funding from the UK Medical Research Council under a concordat with the UK Department for International Development, the NIHR Health Protection Research Unit in Modelling Methodology and Community Jameel. This work used the Cirrus UK National Tier-2 HPC Service at EPCC (http://www.cirrus.ac.uk) funded by the University of Edinburgh and EPSRC (EP/P020267/1). We would like to thank Amazon AWS and Microsoft Azure for computational credits. We would like to thank the Stan Development team for their support. We acknowledge the Medical Research Council and FAPESP (MR/S0195/1).

## Declarations

The authors declare that there is no conflict of interest.

## 6 Supplementary information

### 6.1 Death underreporting scenarios

In this work, an extension of the semi-mechanistic Bayesian hierarchical model from [7] is adopted to reflect the uncertainty about underreported deaths. We address the effect of underreporting in the data set by setting a prior distribution to death underreporting *ψ* ~ *beta*(*θ, ρ*). The hyperparameters of the beta density are fixed in order to reflect in the mode the desired underreporting rate, see Supplementary information Figure 3.

**Figure 3:**
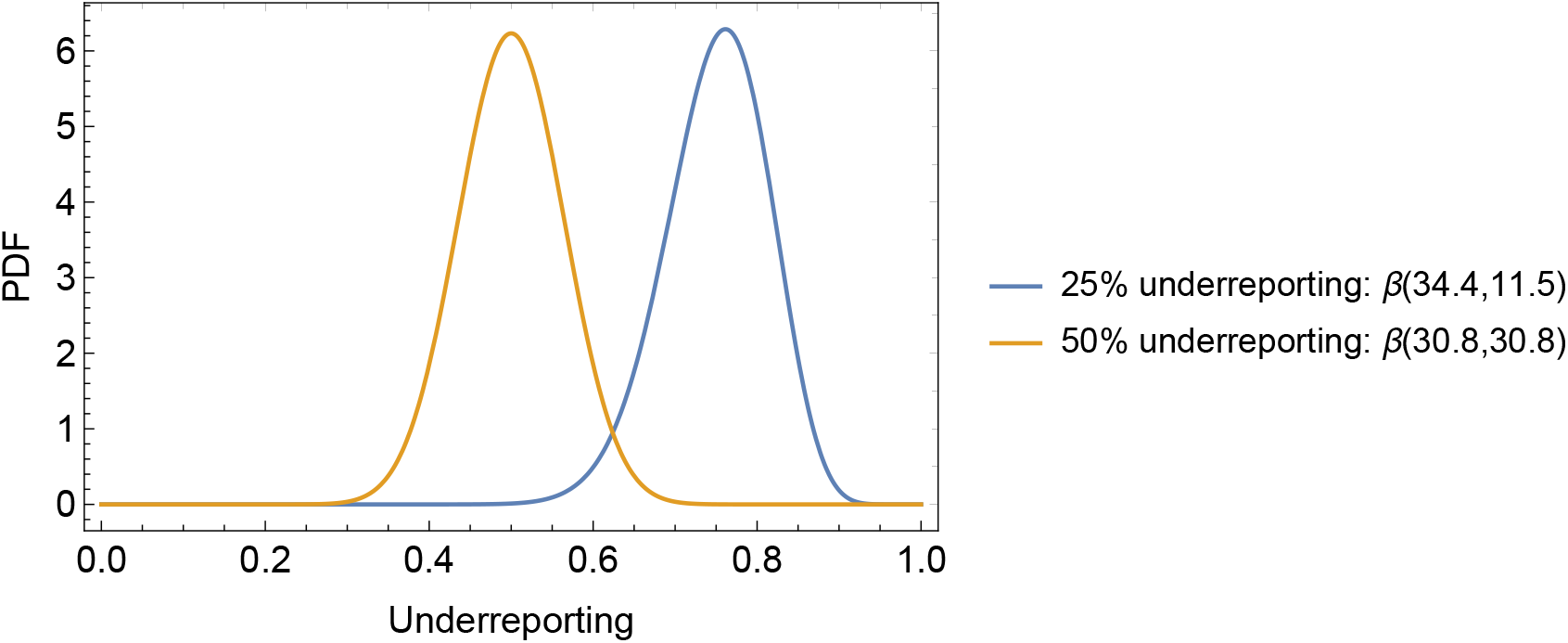
Prior distributions for death underreporting scenarios.

As in the original model [7], daily deaths *D_t_,_m_* are observed for days *t* ∊ {1,…, *n*} and Brazilian states *m* ∊ {1,…,*M*}. These daily deaths are modelled using a positive real-valued function 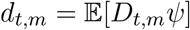 that represents the expected number of deaths attributed to COVID-19, taking into account the designated underreported rate *ψ*. Daily deaths *D_t_,_m_* are assumed to follow a negative binomial distribution with mean *d_t_,_m_* and variance 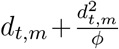, where *ϕ* follows a normal distribution, i.e.

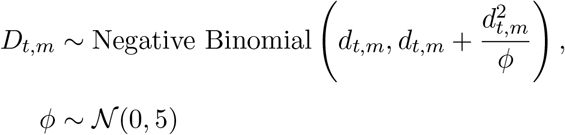

in which 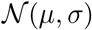 denotes a normal distribution with mean *μ* and standard deviation *σ*. The rest of the mathematical model follows the original manuscript [7] introducing the new feature of underreporting death rate *ψ* on daily deaths.

The effect of death underreporting on the attack rate is shown in Table 2 for three additional scenarios: 50% and 25% and 10% underreporting. The underreporting scenarios are implemented by scaling reported death data by beta distributions with means (0.50, 0.75, 0.90) and in each instance variance 0.004. The distributions are shown in Figure 3.

**Table 2:** Estimated attack rates for 0%, 25% and 50% death underreporting scenarios

| State | 0% underreporting | 25% underreporting | 50% underreporting |
| --- | --- | --- | --- |
| SP | 2.95 [2.05, 4.07] | 4.09 [2.72, 5.81] | 6.07 [3.84, 9.43] |
| RJ | 6.06 [4.47, 7.96] | 8.10 [5.46, 11.80] | 12.30 [7.97, 17.70] |
| PA | 6.09 [4.33, 8.33] | 8.69 [5.25, 13.40] | 12.50 [7.91, 19.40] |
| CE | 5.53 [4.14, 7.20] | 7.33 [5.10, 10.70] | 10.80 [7.29, 15.80] |
| AM | 8.01 [5.73, 10.80] | 10.80 [6.81, 15.40] | 16.50 [10.40, 25.40] |
| PE | 3.01 [2.12, 4.15] | 3.95 [2.54, 5.72] | 6.34 [3.82, 9.73] |
| MA | 1.70 [1.15, 2.41] | 2.35 [1.52, 3.56] | 3.62 [2.15, 5.59] |
| BA | 0.42 [0.26, 0.63] | 0.58 [0.33, 1.00] | 0.84 [0.49, 1.31] |
| ES | 1.41 [0.83, 2.18] | 1.85 [1.07, 3.02] | 2.74 [1.57, 4.54] |
| AL | 1.96 [1.13, 3.08] | 2.64 [1.51, 4.33] | 3.67 [2.06, 6.06] |
| PB | 1.46 [0.87, 2.29] | 2.09 [1.09, 3.67] | 2.87 [1.59, 4.71] |
| MG | 0.14 [0.07, 0.25] | 0.20 [0.10, 0.36] | 0.30 [0.15, 0.54] |
| PR | 0.18 [0.09, 0.32] | 0.23 [0.11, 0.41] | 0.35 [0.17, 0.63] |
| RS | 0.31 [0.15, 0.55] | 0.41 [0.19, 0.77] | 0.68 [0.30, 1.27] |
| RN | 0.61 [0.29, 1.09] | 0.82 [0.35, 1.64] | 1.18 [0.54, 2.14] |
| AP | 3.60 [1.74, 6.34] | 4.55 [2.32, 8.02] | 7.57 [3.57, 14.10] |
| SC | 0.23 [0.09, 0.45] | 0.34 [0.14, 0.68] | 0.41 [0.19, 0.79] |
| GO | 0.30 [0.12, 0.59] | 0.46 [0.19, 0.90] | 0.62 [0.26, 1.17] |

### 6.2 Cases and *R_t_* for 18 states

The estimated cases, deaths and *R_t_* for all 18 states considered in our joint model, São Paulo (SP), Riode Janeiro (RJ), Ceará (CE), Amazonas (AM), Pernambuco (PE), Pará (PA), Maranhão (MA), Bahia (BA), Espírito Santo (ES), Alagoas (AL), Minas Gerais (MG), Paraíba (PB), Paraná (PR), Amapá (AP), Rio Grande do Sul (RS), Rio Grande do Norte (RN), Santa Catarina (SC), Goias (GO), are shown in Figure 4 considering mix of data sets. The same analysis taking into account each data set was also conducted, see Figures 6 and 5.

**Figure 4:**
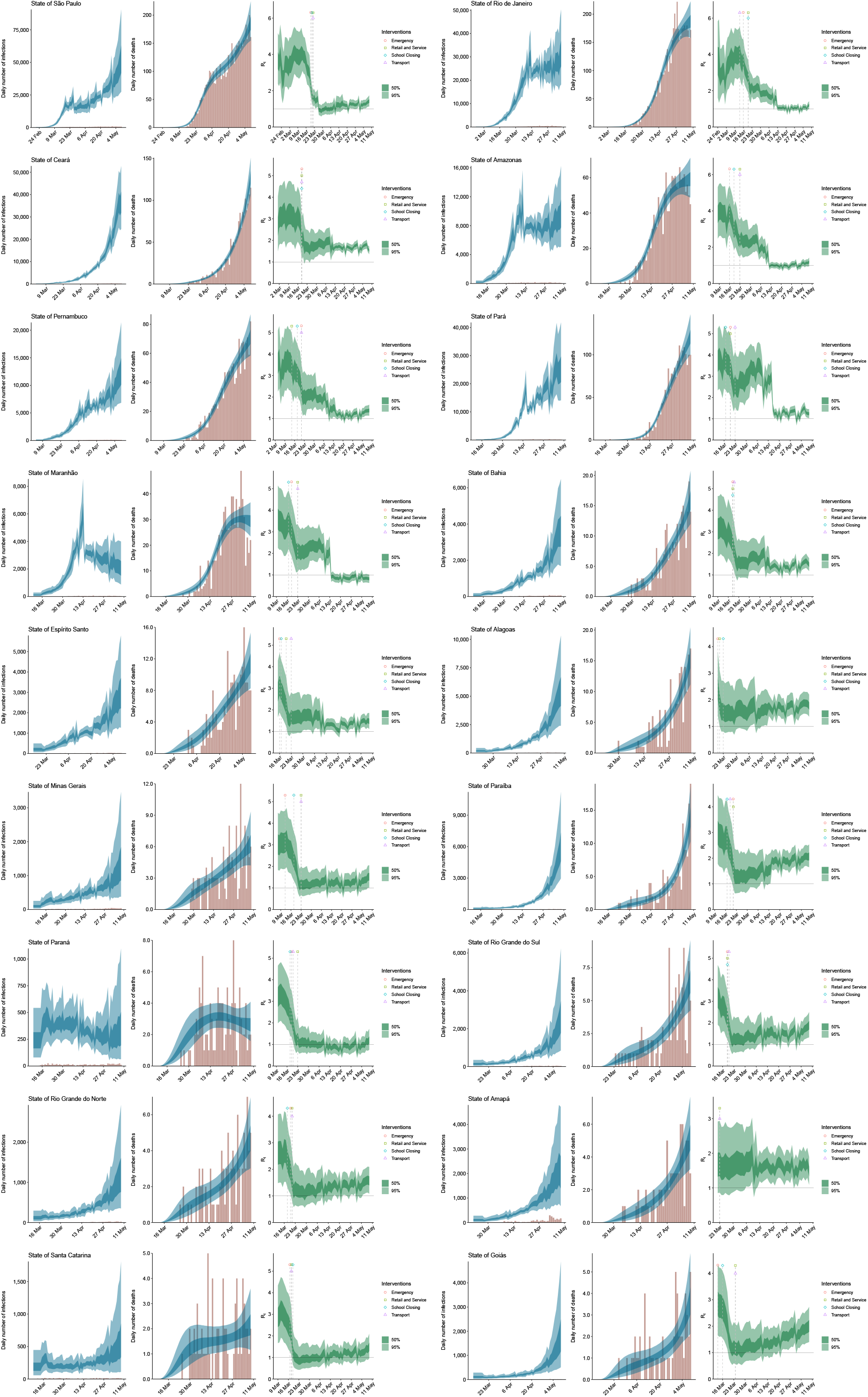
Estimates of infections, deaths and *R_t_* for all 18 states considered in the model considering the mix of both data sets (Painel Coronavírus [19] and SIVEP Gripe[22]).

**Figure 5:**
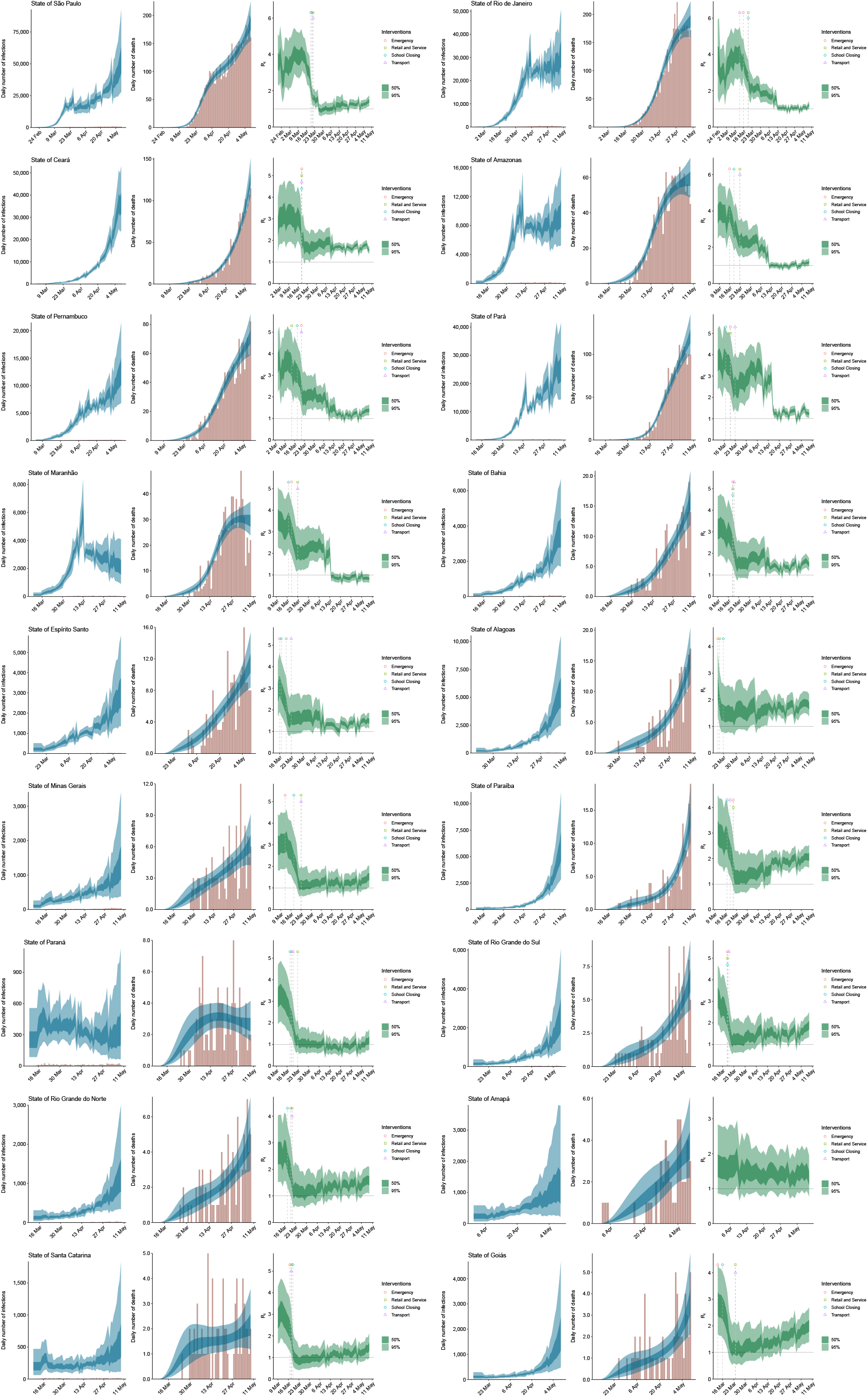
Estimates of infections, deaths and *R_t_* for all 18 states considered in the joint model based on SIVEP-Gripe [22] data.

**Figure 6:**
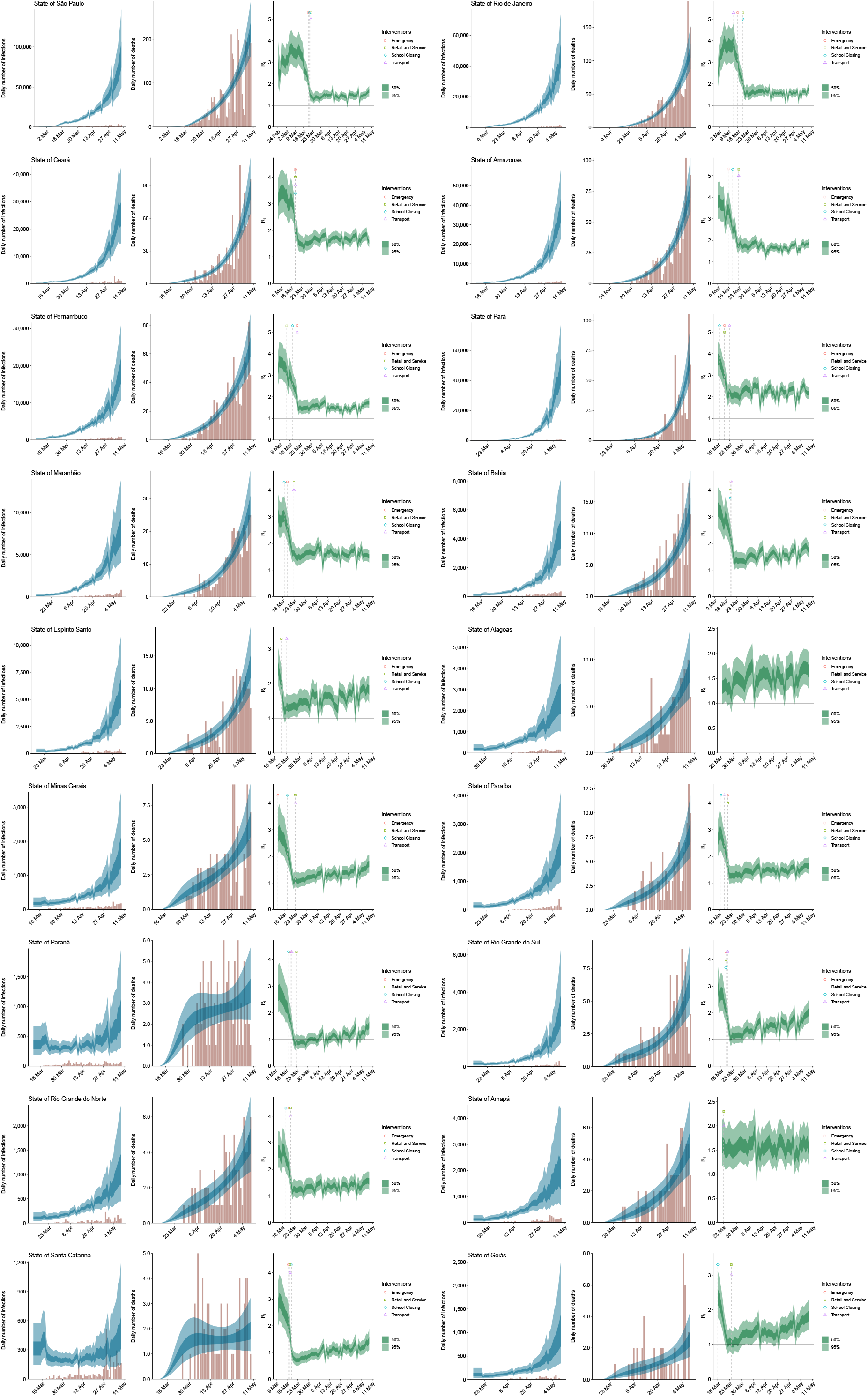
Estimates of infections, deaths and *R_t_* for a joint model of 18 states based on Painel Coronavírus [19] data.

**Figure 7:**
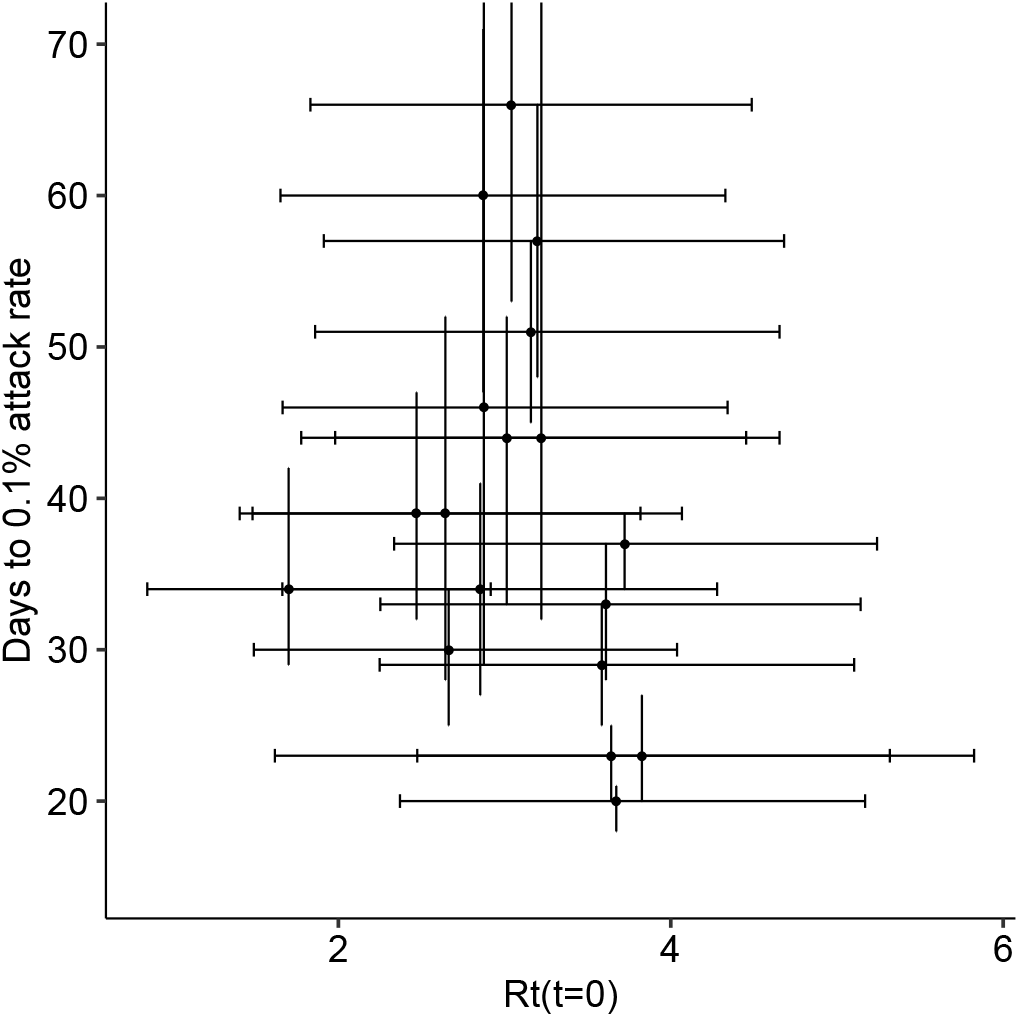
Time in days to 0.1% attack rate (measured from date of first case in Brazil) vs initial reproduction number *R_t_*(*t* = 0) for 18 states considered in the joint model: São Paulo (SP), Rio de Janeiro (RJ), Pará (PA), Ceará (CE), Amazonas (AM), Pernambuco (PE), Maranhão (MA), Bahia (BA), Espárito Santo (ES), Alagoas (AL), Paráába (PB), Minas Gerais (MG), Paraná (PR), Rio Grande do Sul (RS), Rio Grande do Norte (RN), Amapa (AP), Santa Catarina (SC), Goias (GO).

### 6.3 Onset-to-death sensitivity

Onset-to-death sensitivity analysis is shown in Table 3. Attack rates are compared using an onset-to-death gamma distribution fitted specifically to Brazilian data,[22, 11] and an onset-to-death gamma distribution with a longer mean based on earlier estimates.[24] While estimates are quantitatively affected, changes are small in most states, and conclusions overall remained unaltered.

**Table 3:** Attack rate (AR) with onset-to-death distribution for Brazil specifically, r(13.4,0.7) fitted from SIVEP Gripe dataset, and r(17.8,0.45) from [24]

| State | AR% $\Gamma(13.4, 0.7)$ onset-to-death | AR% $\Gamma(17.8, 0.45)$ onset-to-death |
| --- | --- | --- |
| SP | 2.95 [2.05, 4.07] | 3.44 [2.06, 5.42] |
| RJ | 6.06 [4.47, 7.96] | 6.17 [4.09, 8.84] |
| PA | 6.09 [4.33, 8.33] | 6.16 [3.72, 9.60] |
| CE | 5.53 [4.14, 7.20] | 7.55 [4.95, 10.90] |
| AM | 8.01 [5.73, 10.80] | 8.10 [5.22, 12.00] |
| PE | 3.01 [2.12, 4.15] | 3.52 [2.10, 5.53] |
| MA | 1.70 [1.15, 2.41] | 1.63 [1.01, 2.47] |
| BA | 0.42 [0.26, 0.63] | 0.53 [0.27, 0.92] |
| ES | 1.41 [0.83, 2.18] | 1.69 [0.83, 3.00] |
| AL | 1.96 [1.13, 3.08] | 3.09 [1.33, 5.90] |
| PB | 1.46 [0.87, 2.29] | 2.71 [1.24, 5.11] |
| MG | 0.14 [0.07, 0.25] | 0.18 [0.07, 0.37] |
| PR | 0.18 [0.09, 0.32] | 0.19 [0.09, 0.37] |
| RS | 0.31 [0.15, 0.55] | 0.41 [0.15, 0.89] |
| RN | 0.61 [0.29, 1.09] | 0.81 [0.29, 1.78] |
| AP | 3.60 [1.74, 6.34] | 4.85 [1.88, 9.81] |
| SC | 0.23 [0.09, 0.45] | 0.29 [0.10, 0.67] |
| GO | 0.30 [0.12, 0.59] | 0.49 [0.14, 1.16] |

### 6.4 IFR Calculation and Sensitivity Analysis

In order to derive an expected IFR across different states in Brazil, mixing patterns from Latin America [9] and virus’ transmissibility [7] were used. Moreover, to account for the disease severity, data from Chinese epidemic [21] was modified to match data from the outbreak in UK [7]. Additionally we modified these estimates of disease severity (specifically the IFR) to account for the substantial heterogeneity we expect to observe in health outcomes across states due to variation in healthcare quality and capacity, the details of which are described below.

Across the states considered in this analysis, average income (in US dollars) varies from as high as ~ $300 in São Paulo to as low as ~ $100 in Maranhão.[12] Such disparities in income are likely to result in significant differences in the quality and extent of available healthcare. Motivated by this, we modified the state-specific IFRs used in an income-dependent manner. Specifically, we assumed that the state with the highest income (São Paulo) has a quality of care identical to that observed in China (and thus motivated using the estimates presented in Verity et al.[21]), and that the state with the lowest income (Maranhão) had significantly worse healthcare outcomes - more similar to those that would be expected in a Lower Middle Income Country (see [26] for further details on how differences in health quality across settings are likely to impact outcomes). For the other states where income lies somewhere between that of Maranhão and São Paulo, we linearly interpolate the age-specific infection fatality probabilities based on state-level average income.[12] These age-specific infection fatality probabilities are then combined with predictions of the age-distribution of infections to produce an overall, state-specific IFR.

Substantial uncertainty still remains in these IFR calculations. Motivated by this we carried out a sensitivity analysis exploring the impacts of different choices of mixing matrix (Peru vs the United Kingdom) and of assumptions surrounding healthcare quality (namely the interpolation method described above or assuming that all states are able to provide a level of healthcare equal to that seen during the Chinese epidemic). The results of these sensitivity analyses are shown in Table 4 for different IFRs. Although assumptions surrounding healthcare quality impact the quantitative predictions of the IFR and associated predicted attack rates, they do not qualitatively change our conclusions surrounding herd immunity and the lack of infections to date sufficient to have reached it.

**Table 4:** Attack rates % (AR) estimated using different infection fatality ratios (IFR) with Brazilian state-level population weighting and using: i) UK contact matrix, ii) Peru contact matrix, iii) UK contact matrix with poorer hospitalisation outcomes, iv) Peru contact matrix with poorer hospitalisation outcomes.

| State | (i) AR% UK con-<br>tact matrix | (ii) AR% Peru con-<br>tact matrix | (iii) AR% UK con-<br>tact matrix, poorer<br>outcomes | (iv) AR% Peru<br>contact matrix,<br>poorer outcomes |
| --- | --- | --- | --- | --- |
| SP | 3.07 [2.14, 4.21] | 2.95 [2.06, 4.07] | 3.07 [2.14, 4.22] | 2.95 [2.05, 4.07] |
| RJ | 6.62 [4.89, 8.72] | 6.33 [4.68, 8.32] | 6.31 [4.68, 8.28] | 6.06 [4.47, 7.96] |
| PA | 12.60 [9.04, 17.10] | 12.30 [8.85, 16.70] | 6.11 [4.32, 8.32] | 6.09 [4.33, 8.33] |
| CE | 10.20 [7.63, 13.20] | 9.73 [7.34, 12.70] | 5.63 [4.19, 7.36] | 5.53 [4.14, 7.20] |
| AM | 16.70 [12.10, 22.30] | 16.50 [12.00, 22.10] | 7.97 [5.70, 10.80] | 8.01 [5.73, 10.80] |
| PE | 5.54 [3.92, 7.57] | 5.33 [3.74, 7.34] | 3.06 [2.16, 4.19] | 3.01 [2.12, 4.15] |
| MA | 3.70 [2.49, 5.23] | 3.57 [2.41, 5.04] | 1.73 [1.17, 2.44] | 1.70 [1.15, 2.41] |
| BA | 0.80 [0.49, 1.19] | 0.76 [0.48, 1.13] | 0.43 [0.27, 0.64] | 0.42 [0.26, 0.63] |
| ES | 2.00 [1.20, 3.09] | 1.92 [1.15, 2.97] | 1.44 [0.86, 2.24] | 1.41 [0.83, 2.18] |
| AL | 4.02 [2.37, 6.30] | 3.90 [2.28, 6.11] | 1.96 [1.15, 3.11] | 1.96 [1.13, 3.08] |
| PB | 2.68 [1.58, 4.17] | 2.55 [1.51, 3.97] | 1.49 [0.89, 2.33] | 1.46 [0.87, 2.29] |
| MG | 0.22 [0.11, 0.38] | 0.21 [0.10, 0.36] | 0.15 [0.07, 0.26] | 0.14 [0.07, 0.25] |
| PR | 0.24 [0.12, 0.42] | 0.23 [0.11, 0.40] | 0.19 [0.09, 0.33] | 0.18 [0.09, 0.32] |
| RS | 0.35 [0.17, 0.62] | 0.33 [0.16, 0.58] | 0.32 [0.16, 0.57] | 0.31 [0.15, 0.55] |
| RN | 1.08 [0.51, 1.94] | 1.03 [0.48, 1.87] | 0.63 [0.29, 1.12] | 0.61 [0.29, 1.09] |
| AP | 7.70 [3.82, 13.50] | 7.52 [3.70, 13.10] | 3.60 [1.75, 6.34] | 3.60 [1.74, 6.34] |
| SC | 0.28 [0.12, 0.53] | 0.27 [0.11, 0.51] | 0.24 [0.10, 0.46] | 0.23 [0.09, 0.45] |
| GO | 0.48 [0.20, 0.92] | 0.47 [0.20, 0.91] | 0.30 [0.12, 0.59] | 0.30 [0.12, 0.59] |

**Table 5:** Infection fatality ratios (IFR) with Brazilian state-level population weighting,, using: i) UK contact matrix, ii) Peru contact matrix, iii) UK contact matrix with poorer hospitalisation outcomes, iv) Peru contact matrix with poorer hospitalisation outcomes.

| State | (i) IFR UK contact matrix | (ii) IFR Peru contact matrix | (iii) IFR UK contact matrix, poorer outcomes | (iv) IFR Peru contact matrix, poorer outcomes |
| --- | --- | --- | --- | --- |
| AC | 0.38 | 0.39 | 0.78 | 0.78 |
| AL | 0.51 | 0.53 | 1.06 | 1.07 |
| AM | 0.37 | 0.38 | 0.79 | 0.79 |
| AP | 0.34 | 0.35 | 0.73 | 0.73 |
| BA | 0.59 | 0.62 | 1.10 | 1.12 |
| CE | 0.58 | 0.61 | 1.07 | 1.09 |
| ES | 0.63 | 0.65 | 0.87 | 0.89 |
| MA | 0.48 | 0.50 | 1.03 | 1.04 |
| MG | 0.69 | 0.72 | 1.01 | 1.04 |
| PA | 0.42 | 0.43 | 0.89 | 0.89 |
| PB | 0.62 | 0.65 | 1.13 | 1.16 |
| PE | 0.58 | 0.60 | 1.06 | 1.07 |
| PI | 0.57 | 0.59 | 1.10 | 1.11 |
| PR | 0.66 | 0.69 | 0.84 | 0.86 |
| RJ | 0.73 | 0.76 | 0.76 | 0.79 |
| RN | 0.60 | 0.62 | 1.04 | 1.06 |
| RO | 0.45 | 0.45 | 0.81 | 0.81 |
| RR | 0.33 | 0.34 | 0.67 | 0.67 |
| RS | 0.78 | 0.81 | 0.84 | 0.87 |
| SC | 0.65 | 0.67 | 0.74 | 0.76 |
| SE | 0.51 | 0.53 | 0.96 | 0.97 |
| SP | 0.67 | 0.70 | 0.67 | 0.70 |
| TO | 0.49 | 0.51 | 0.89 | 0.90 |

**Table 6:**
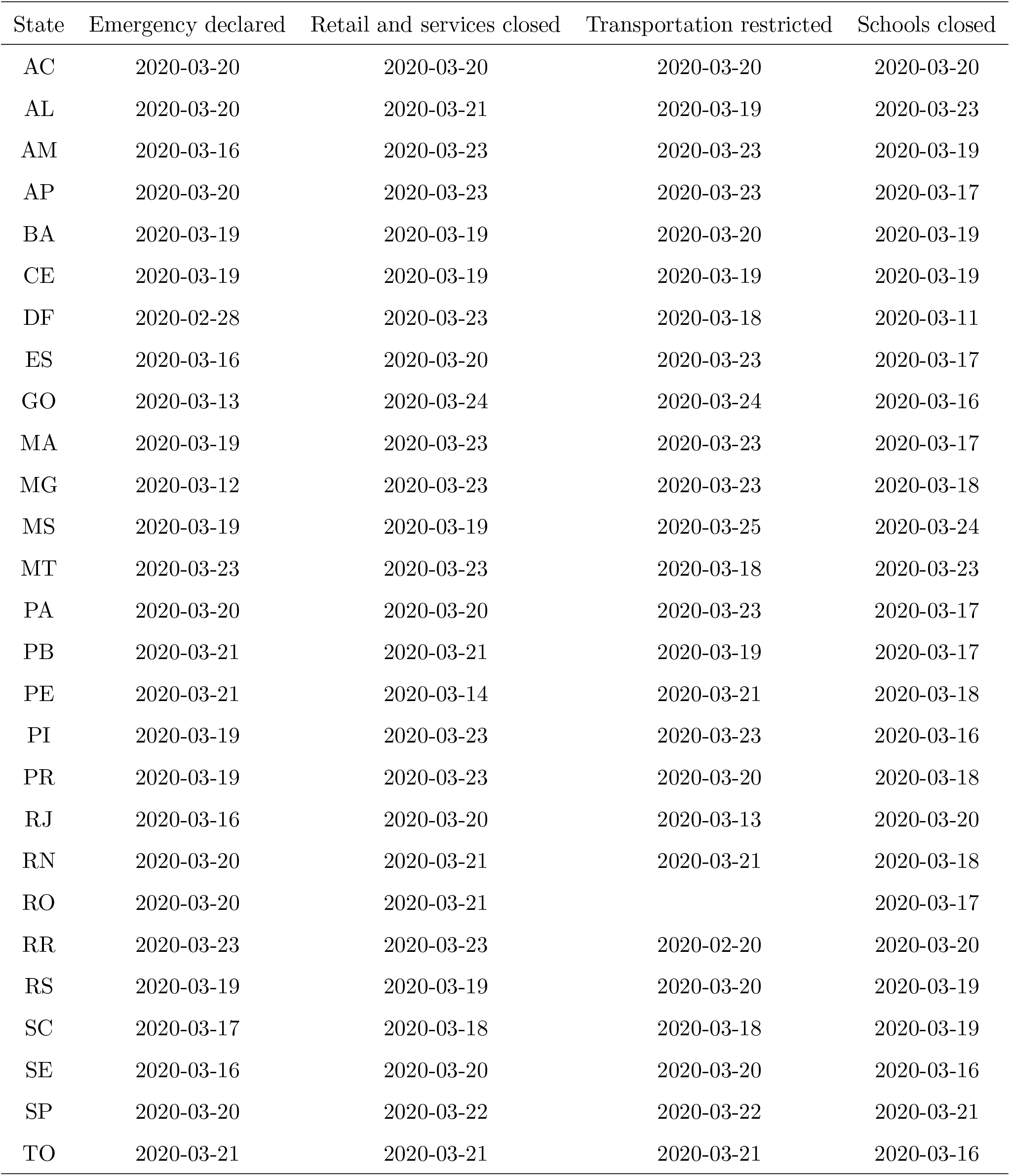
Non-pharmaceutical interventions by state, adapted from [3].

| State | Emergency declared | Retail and services closed | Transportation restricted | Schools closed |
| --- | --- | --- | --- | --- |
| AC | 2020-03-20 | 2020-03-20 | 2020-03-20 | 2020-03-20 |
| AL | 2020-03-20 | 2020-03-21 | 2020-03-19 | 2020-03-23 |
| AM | 2020-03-16 | 2020-03-23 | 2020-03-23 | 2020-03-19 |
| AP | 2020-03-20 | 2020-03-23 | 2020-03-23 | 2020-03-17 |
| BA | 2020-03-19 | 2020-03-19 | 2020-03-20 | 2020-03-19 |
| CE | 2020-03-19 | 2020-03-19 | 2020-03-19 | 2020-03-19 |
| DF | 2020-02-28 | 2020-03-23 | 2020-03-18 | 2020-03-11 |
| ES | 2020-03-16 | 2020-03-20 | 2020-03-23 | 2020-03-17 |
| GO | 2020-03-13 | 2020-03-24 | 2020-03-24 | 2020-03-16 |
| MA | 2020-03-19 | 2020-03-23 | 2020-03-23 | 2020-03-17 |
| MG | 2020-03-12 | 2020-03-23 | 2020-03-23 | 2020-03-18 |
| MS | 2020-03-19 | 2020-03-19 | 2020-03-25 | 2020-03-24 |
| MT | 2020-03-23 | 2020-03-23 | 2020-03-18 | 2020-03-23 |
| PA | 2020-03-20 | 2020-03-20 | 2020-03-23 | 2020-03-17 |
| PB | 2020-03-21 | 2020-03-21 | 2020-03-19 | 2020-03-17 |
| PE | 2020-03-21 | 2020-03-14 | 2020-03-21 | 2020-03-18 |
| PI | 2020-03-19 | 2020-03-23 | 2020-03-23 | 2020-03-16 |
| PR | 2020-03-19 | 2020-03-23 | 2020-03-20 | 2020-03-18 |
| RJ | 2020-03-16 | 2020-03-20 | 2020-03-13 | 2020-03-20 |
| RN | 2020-03-20 | 2020-03-21 | 2020-03-21 | 2020-03-18 |
| RO | 2020-03-20 | 2020-03-21 |  | 2020-03-17 |
| RR | 2020-03-23 | 2020-03-23 | 2020-02-20 | 2020-03-20 |
| RS | 2020-03-19 | 2020-03-19 | 2020-03-20 | 2020-03-19 |
| SC | 2020-03-17 | 2020-03-18 | 2020-03-18 | 2020-03-19 |
| SE | 2020-03-16 | 2020-03-20 | 2020-03-20 | 2020-03-16 |
| SP | 2020-03-20 | 2020-03-22 | 2020-03-22 | 2020-03-21 |
| TO | 2020-03-21 | 2020-03-21 | 2020-03-21 | 2020-03-16 |

### 6.5 Death data source interpretation and auxiliary model data

As deaths are only reported if SARS-CoV-2 is confirmed by a positive result on diagnostic testing, there can be considerable and variable lag between the date of death and the official reporting of COVID-19 deaths. Current Ministry of Health protocol guides that all hospitalizations and deaths due to suspected or confirmed COVID-19 must also be notified on an online system called SIVEP-Gripe.[22] After testing and investigation by health officials, cases and deaths receive a final COVID-19 classification and records are updated to include lab results and the exact date of death or hospital discharge for each patient. Thus, SIVEP-Gripe allows for the epidemic to be tracked by the actual daily number of COVID-19 deaths. For instance, the official *Painel Coronavírus* daily death count on the 9th of May 2020 was 732, but there are at least 799 COVID19-confirmed deaths already registered on SIVEP-Gripe. In total, SIVEP-Gripe records 18,314 confirmed COVID-19 deaths, 72.3% more than the official 10,627 count for the same date. However, as not all states are following the official protocol of recording their patients on SIVEP-Gripe in a timely manner (see Figure 8), the true number of COVID-19 deaths on any given day can be significantly larger.

**Figure 8:**
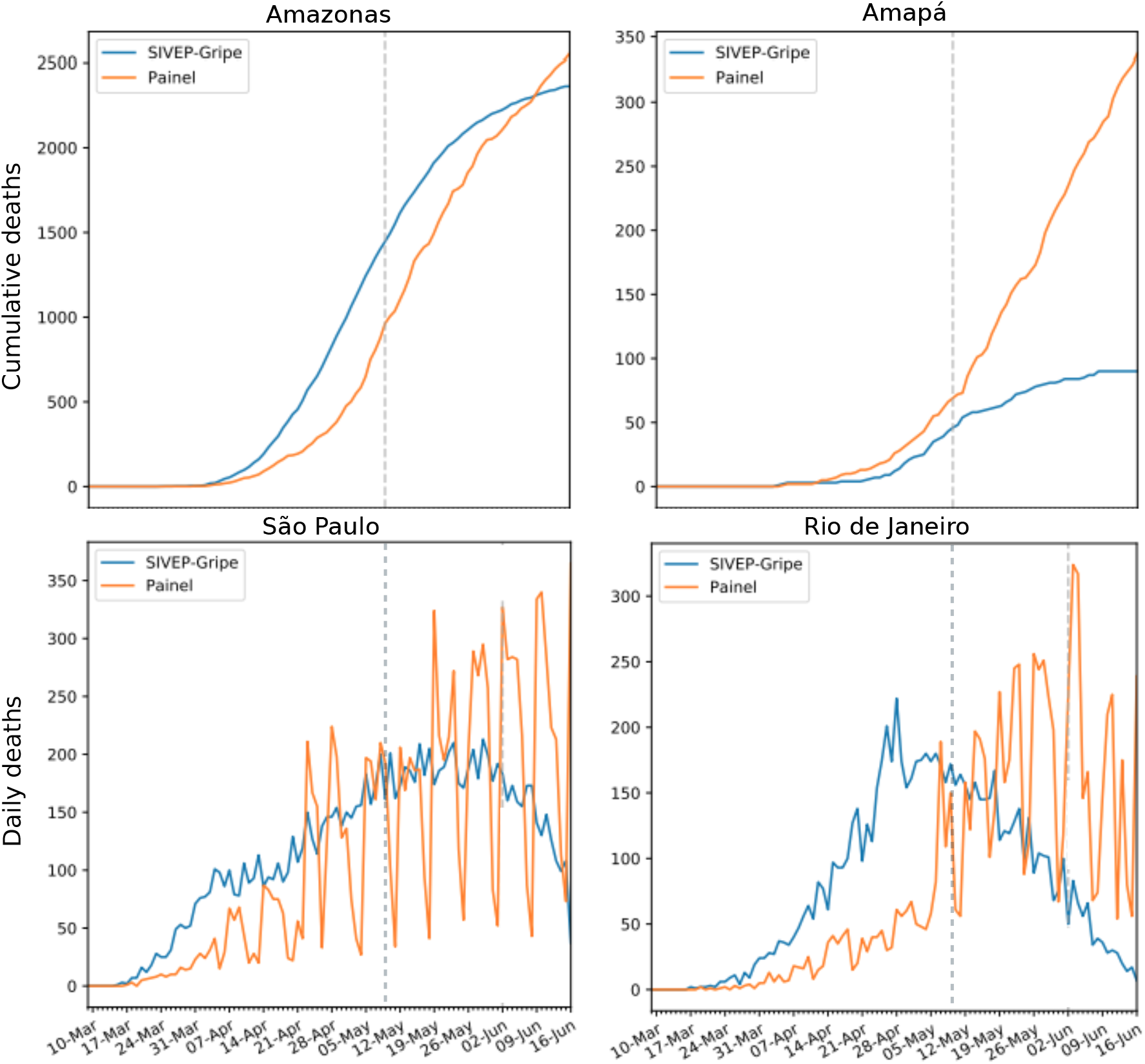
Comparison of the daily death records from Brazil’s Ministry of Health *Painel Coronavírus* (date of notification - orange line)[19] and SIVEP-Gripe (date of death - blue line)[22] datasets, from 10 March 2020 to 16 June 2020. Gray dashed vertical line represents 09 May, the last day in our analysis. Top row displays cumulative deaths plots. Top left, state of Amazonas (population 4.2M), shows a lag of approximately 10 days between the cumulative counts of deaths by date of death vs. date of notification. Top right, state of Amapá (population 861,000), has the cumulative count of deaths by date of notification higher than by date of death, indicating deaths are being reported to the Ministry of Health but not updated onto the SIVEP-Gripe database. Bottom row are daily death plots. Bottom left, state of São Paulo (population 46M), shows the effects of processing and testing delays on the official daily counts, and also the uncertain effect of right-censoring. Bottom right, state of Rio de Janeiro (population 17M), shows testing and processing lags. Important to caution that there is still substantial, but uncertain, right-censoring in the SIVEP-Gripe data for the state of Rio de Janeiro.

**Figure 9:**
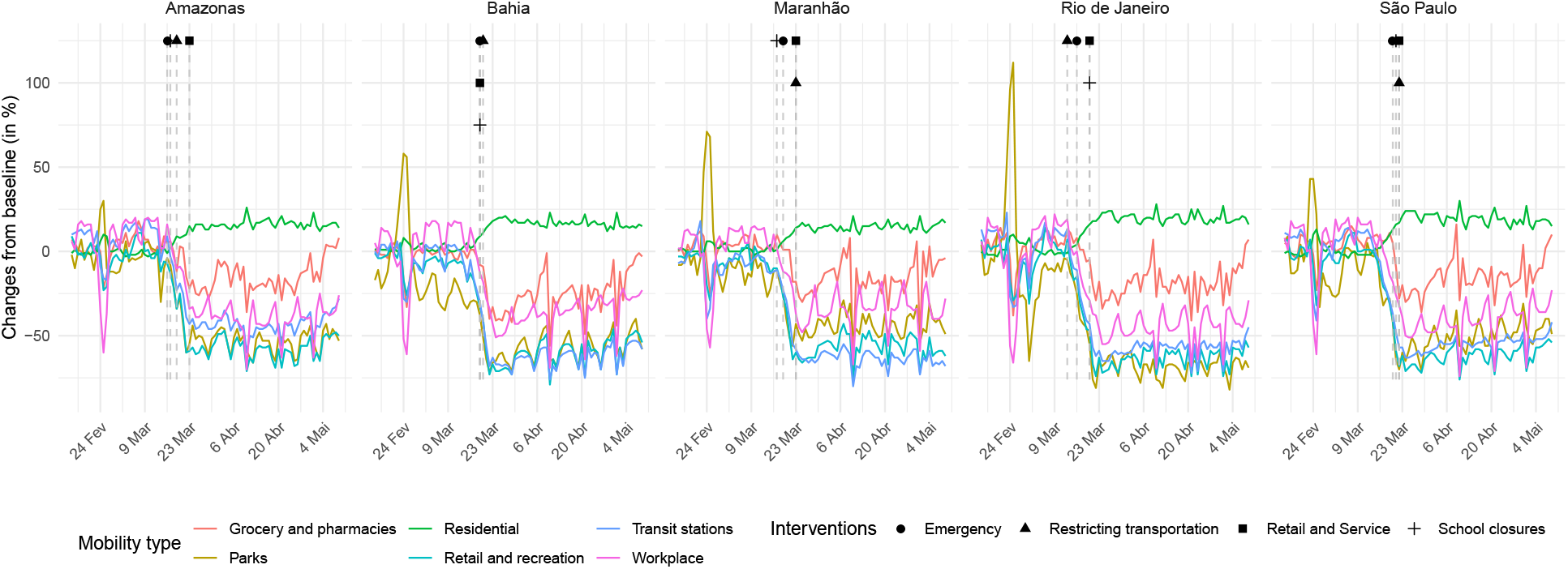
Mobility covariates from Google mobility reports for São Paulo (SP), Rio de Janeiro (RJ), Pernambuco (PE), Ceará (CE), Amazonas (AM).

For population counts we used the 2020 projection by state published by *Instituto Brasileiro de Geografia e Estatística* (IBGE).[13]

Mobility report data from Google (https://www.google.com/covid19/mobility/) were used to estimate the effects of different interventions over time. The report provides the estimated percentage of change on movements of places such as retail and recreation, groceries and pharmacies, parks, transit stations, workplaces, and residential comparing to a baseline. Such baseline corresponds to the median value of each day of the week, using data of January 3rd to February 6th, 2020.

Regarding intervention data, the values taken into account are the dates in which interventions were effectively applied, even though they were encouraged at earlier dates.

## Notes

### Competing Interest Statement

The authors have declared no competing interest.

### Author Declarations

This study is based on data available in the public domain.

### Summary of Updates

Anlaysis updated to include the data from the SRAG database. Model updated to include autogressive component.

